# Quantitative Assessment of Injectable Medication Delivery Practices

**DOI:** 10.1101/2023.01.09.23284320

**Authors:** Salena Aggerwal, Amir Minerbi, Lt(N) Peter Beliveau, LCol Sean Meredith, MCpl Sasha Lalonde, Erica Laurin, Gaurav Gupta

## Abstract

**Background:** While medical advances for in-hospital care rapidly evolve, a mainstay of effective pre-hospital care remains the ability to treat medical emergencies such as anaphylaxis, overdosing, and/or uncontrolled bleeding through rapid administration of appropriate medication. Therefore, investigators looked at various injection methods and their possible utility in medical emergencies.

**Method:** 30 participants were asked to inject ‘medication’ that mimicked three different methods of injection: 1) autoinjectors, 2) prefilled syringes, and 3) traditional standard syringes using clinical scenarios. Three variables that were measured in the study were: the time required to complete the injection, the perceived difficulties, and the participant’s performance errors.

**Results:** The perceived difficulty and injection time for the autoinjector device were statistically significantly lower compared to prefilled syringes and standard syringes. No significant difference in errors were seen between platforms.

**Discussion:** To our knowledge, this is the first study to quantify the gain of efficiency when comparing autoinjectors to other methods of medication administration, like prefilled syringes or drawing medication from vials for administration. The clinical implications of the noted differences are not clear at this time. Many potential limitations exist, including the size of the study, the use of non-clinical participants, the immediate use of platforms after training, and the lack of applied stress in the environment.

**Conclusion:** This study compares autoinjectors to other methods of medication administration; prefilled syringes and standard syringes. Further study in larger datasets with clinicians and/or military personnel is required to compare these platforms in various environments. The outcome of this project provides insights into the relative efficiencies of treating medical emergencies such as anaphylaxis, overdosing, and/or uncontrolled bleeding.

## Background

The mainstay of effective pre-hospital care is the ability to treat medical emergencies such as anaphylaxis, overdosing, and/or uncontrolled bleeding, through rapid administration of appropriate medication. Rapid administration of intramuscular medications are essential when oral or intravenous delivery is either not possible or ineffective, often in environments where immediate access to medical facilities are limited.

Medical teams and civilians worldwide carry multiple emergency autoinjectors and drug kits manufactured and purchased from various sources (e.g. DuoDote, EpiPen, Morphine, Benzodiazepines, Tranexamic Acid, biological warfare antidotes, Narcan, neuroleptics) (1-3). Nearly all autoinjectors and drug kits have different usage instructions, expiry dates, indications, and sizes, and to this point, have been restricted and manufactured for a single drug. Prior studies of such autoinjectors in various settings have shown that the variability across the devices are factors that make injectable drug delivery error-prone, cumbersome, confusing, bulky, and slow (4-9). However, there is also no data that we are aware of that quantifies the gain of efficiency when comparing autoinjectors to other methods of medication administration like prefilled syringes, or drawing medication from vials for administration.

As a result, there is a need to improve upon the currently available syringe, needle, and injector systems. There should be a focus on improved medication delivery systems that prioritizes increased speed, ease of use, versatility, and reducing administration errors. For tactical medical care, there is also a need to reduce equipment bulk and volume, to improve the efficiency of these tools, and to reduce cognitive loads on the providers. Solutions include decoupling the injectable medication from the auto-injector itself, allowing a single needle to be used with various medications. This is instead of having multiple autoinjectors for various drugs, as is the current standard in the auto-injector market (i.e. Epipen). The other option includes creating adapters for existing syringe and needle systems that facilitate medication delivery. The proposed benefits of this device include making drug delivery quicker, safer, and more cost-effective.

The intent of this study is to understand the efficiency of injectable medication and to identify areas of potential improvement. The outcome from this work could improve the safety and efficacy of emergency medical services, by developing medication delivery devices that will reduce physical carrying load, improve response time for life-saving interventions, as well as improve the comfort and safety of injectable medications for end users. Specifically, we aim to determine the efficiency of medication injection when comparing various strategies.

## Methods

Between April and June 2022, 30 participants completed the study. Inclusion criteria consisted of non-healthcare professionals, including males and females between the age of 18-60, with no experience with medication injection during the last 2 years. Exclusion criteria were as follows: injuries to participants’ hands (i.e. significant bruising, swollen hands, fractured fingers), cognitive problems (i.e. concussions), and/or drug and alcohol intoxication. Non healthcare professionals were used to establish a baseline performance for medication administration not influenced by experience and to simulate an emergency situation where trained support was not present. The study was approved by the Defence Research Ethics Board Toronto, and all participants gave consent to participate(?).

For each scenario participants were asked to inject the “medication” using three methods represented in Figure 1: 1) standard protocol, 2) autoinjector, and 3) prefilled syringes. For each scenario, the appropriate medication administration type was selected amongst groups of options based on the given instructions and scenario: 1) autoinjector equivalents (i.e. medication, needle and syringe attached); 2) prefilled syringes (i.e. a needle is attached by the participant prior to administration); and 3) standard protocol (i.e. drawing medication from the vial and injecting via syringe and needle). Medication administration scenarios were for either 1) Naloxone (opioid overdose), 2) Epinephrine (anaphylaxis), or 3) Tranexamic acid (bleeding). A 21-gauge 1.5-inch needle was used for drawing up the medication and for injection.

**Figure 1:**
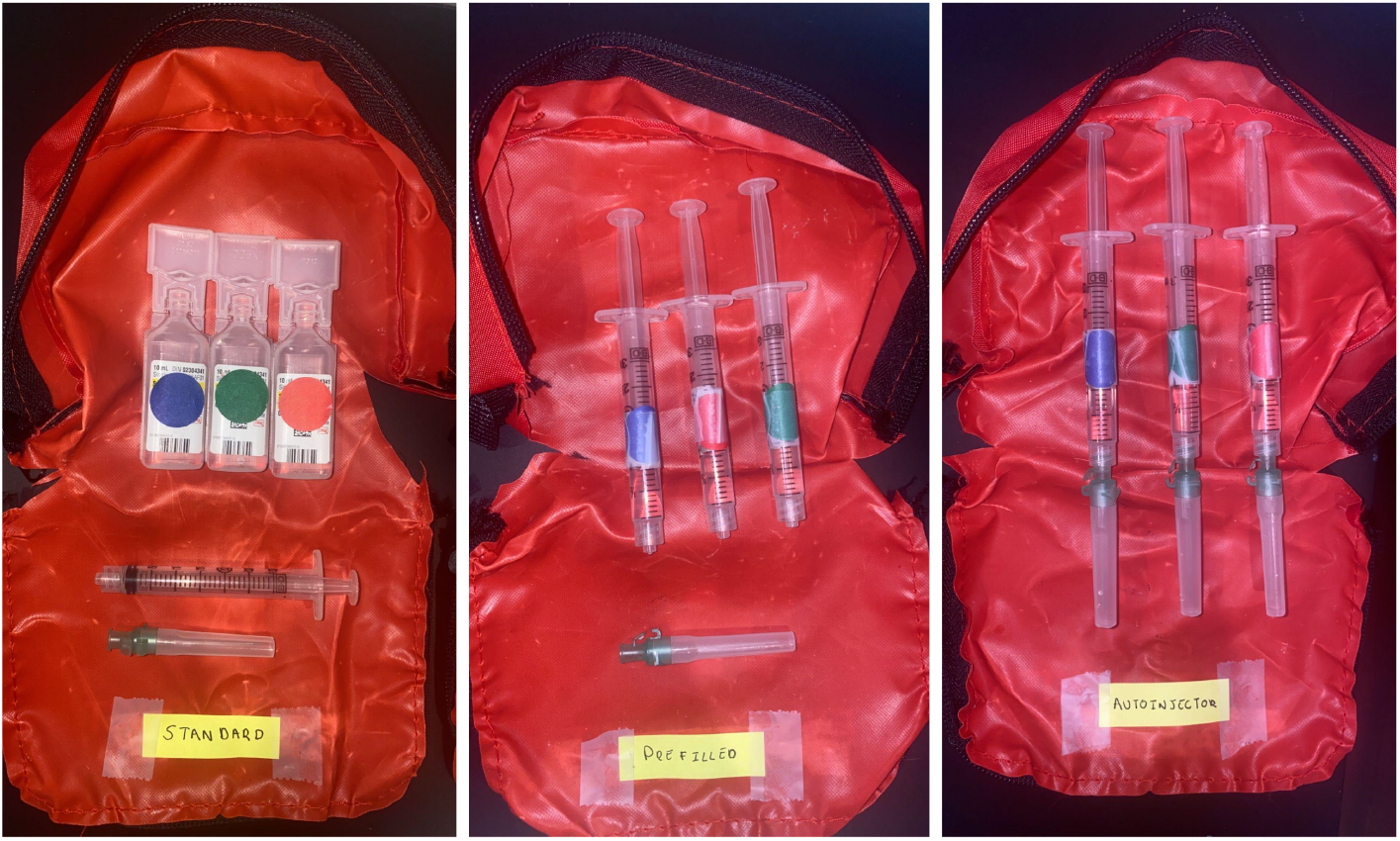
The Three Methods of Injection (i.e. Standard, Prefilled and Autoinjector)

At the end of the experiment, participants were asked how difficult they found each method. Accordingly, they ranked each method on a scale from 1-5, one being the easiest and five being the hardest. The order of administration for each scenario and injection type was randomized to exclude any bias. To minimize participant risk, sterile water was used as a medication substitute, therefore active drug products and live/loaded autoinjectors were not used. The target injections were placed in an inanimate object (i.e. orange) to simulate an injection experience. Time was evaluated using a stopwatch and started at the call of the examiner. Time was stopped once the medication had been delivered.

Any human errors or device failures were recorded. An error was noted if a participant used the wrong injection system or administered the wrong medication for a given scenario.

Each injection method trial was done 3 times each using various scenarios for a total of 9 trials. Before beginning the timed trial, all participants were given the same three practice trials in the same order, which allowed them to try every method of injection.

## Results

Thirty participants were enrolled and completed the study. Participants were mostly women (70%), their mean age was 35.8 (± 16.9) years, and 93% were right-handed. Normality of distribution was tested for injection time and perceived difficulty using Shapiro-Wilk test, and both showed non-normal distribution.

The non-parametric Kruskal-Wallis test was used to compare the means of these variables. Injection times were significantly shorter for autoinjector as compared to both standard syringes and prefilled syringes (Table 2, Figure 2). Statistical differences were also noted when comparing standard to prefilled syringes. When medications were administered via autoinjectors, it was 3.53 and 14.71 seconds faster than the prefilled and standard syringes, respectively. Administration of prefilled syringes were 11.72 seconds faster than the standard syringes. When considering the change in injection time over trials, there was a trend towards shorter injection time in the second and third trials, but it did not reach statistical significance (Friedman’s repeated measure ANOVA p=0.497). Perceived difficulty of injection was significantly lower for the autoinjector compared to standard and prefilled syringes, and prefilled syringes when compared to standard syringes (Table 3, Figure 3). Error rates were comparable in all three methods (Table 3).

**Table 1:** Demographics (Population Descriptive Statistics)

| Variable | Value (N=30) |  |
| --- | --- | --- |
| Average Age | 35.8 yrs |  |
| Sex assigned at birth (F:M) | 70% | 30% |
| Dominant Hand (Left:Right) | 6.7% | 93.3% |

**Table 2:** Injection time (s). Values are presented as mean (standard deviation)

|  | Trial 1<br>(90) | Trial 2<br>(90) | Trial 3<br>(90) | Mean 3<br>trials (270) |
| --- | --- | --- | --- | --- |
| Standard syringe | 24.71<br>(5.17) | 22.77<br>(5.32) | 21.20<br>(4.96) | 22.89<br>(5.29) |
| Prefilled syringe | 12.43<br>(3.82) | 11.80<br>(3.19) | 10.89<br>(2.63) | 11.71<br>(3.28) |
| Autoinjector | 9.33 (4.47) | 7.53 (2.00) | 7.69 (3.55) | 8.18 (3.55) |

**Figure 2:**
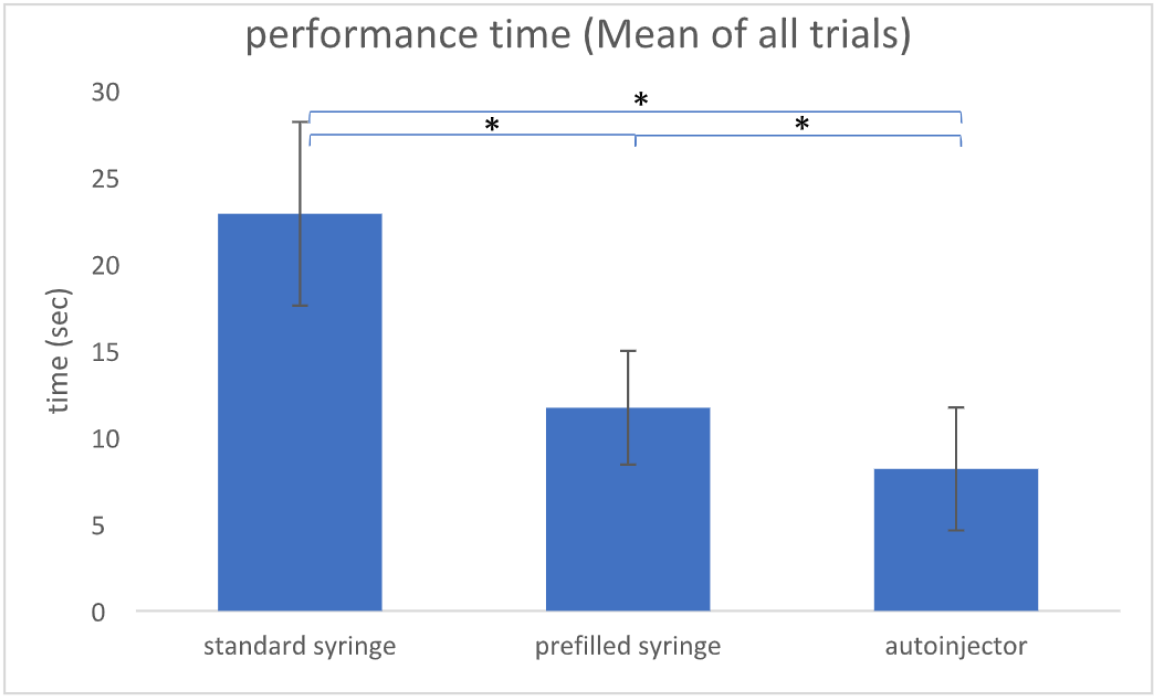
Performance time per injection method, mean of all trials (* p<0.0001)

**Table 3:**
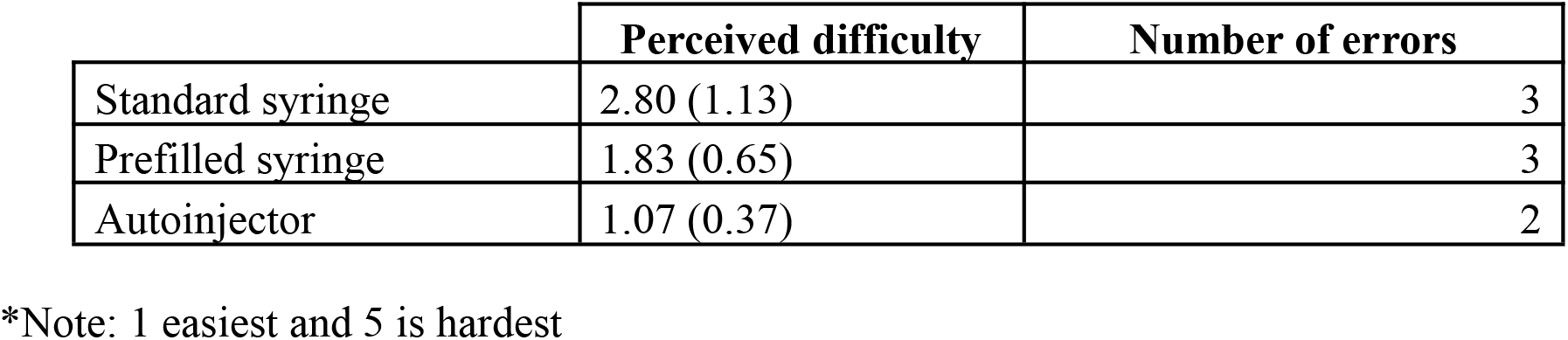
Ease of performance and error rate. Values are presented as mean (standard deviation).

**Figure 3:**
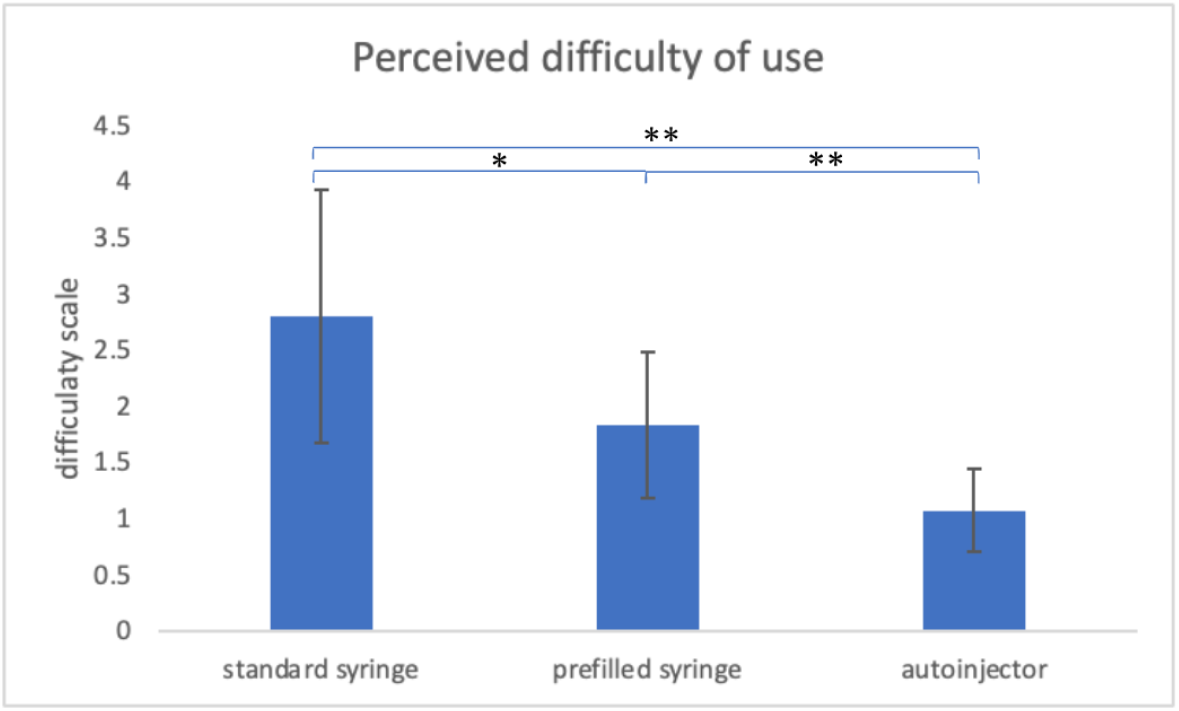
Perceived difficulty of use per injection method (* p<0.05, **p<0.001)

### Statistical Analysis

Quantitative variables were evaluated for normality of distribution using the Wilk-Shapiro test. Normally distributed variables were compared using ANOVA while non-normally distributed variables were compared using the non-parametric Kruskal-Wallis test. Adjustment for multiple comparisons was performed using Bonferroni correction. Non-parametric repeated measure comparison was performed using Friedman’s repeated measures ANOVA. Analyses were done on IBM SPSS version 28.

## Discussion

To our knowledge, this is the first study that aims to quantify the gain of efficiency when comparing autoinjectors to other methods of medication administration, like prefilled syringes or drawing medication from vials for administration.

According to previous research, autoinjectors are seen to be advantageous over other methods with respect to patient/provider safety, human error and cognitive load. Their construction also allows for convenience and easier administration, which may help improve dose accuracy and efficiency. This also helps to benefit patients that may need to self-administer medication as it would improve their self-confidence, reduce their anxiety, and promote compliance [11].

Like autoinjectors, prefilled syringes have advantages over standard vials and syringes, including less overfilling, which can lead up to 20-30% of potential waste in medication [14]. Prefilled syringes could therefore minimize drug wastage, specific platforms may have a longer shelf life, and proponents argue for greater ease of use [14].

In this study both autoinjectors and prefilled devices yielded statistically different results from standard methods when considering efficiency and perceived difficulty of medication administration. Therefore, the clinical utility requires further investigation when compared to the standard method, however the clinical relevance of the associated average difference is not clear at this time.

Many potential limitations exist for this study. Variables not accounted for in this study include the impact of stress on medication administration such as utilization in austere environments, extreme temperatures, and low light situations, waste generated by the different injection methods, and specific measures of cognitive and physical abilities on performance with various platforms. Further study into the impact of various injection methods on various injection methods on age (i.e. the pediatric and elderly population) could be considered as well (11-13). Another limitation was the use of inanimate objects, which is arguably different from human beings. Using a 21-gauge needle for injection compared to a thinner needle, for instance, may be more painful for real patients. Therefore, not switching to a lower gauge could have inflated the efficiency of the standard method. This project also did not account for the recall over time since participants immediately entered the study after training. If time between instruction and administration were lengthened, it is possible that “easier-to-use” platforms (e.g. autoinjectors) may have had a significant effect size related outcome. Finally, this was done with participants without recent injection experience, whereas perhaps the inclusion of those with military and/or clinical backgrounds could have produced different results.

## Conclusions

Autoinjector devices have proven to be significantly more efficient in comparison to prefilled and current traditional methods of medication administration. Prefilled methods have also shown efficiency benefits over drawing from vials and syringes in this trial. Further analysis with larger datasets, participants with clinical/military training, and under stressful situations would be useful before concluding the utility of changing current methods. This project could potentially improve the quality of pre-hospital care in medical emergencies. Further work to control confounders, reported outcomes, and limitations will be required to fully determine the utility of using prefilled devices in medical emergencies, such as anaphylaxis, overdosing, and uncontrolled bleeding.

## Data Availability

All data produced in the present work are contained in the manuscript

## Acknowledgements

Authors would like to thank Aidan McParland from the University of British Columbia for their contribution on protocol development.

## References

1. Institute of Medicine (US) Committee to Study the Interactions of Drugs, Biologics, and Chemicals in U.S. Military Forces; Petersdorf RG, Page WF, Thaul S, editors. Interactions of Drugs, Biologics, and Chemicals in U.S. Military Forces. Washington (DC): National Academies Press (US); 1996. 2, Current Prophylactic Agents. Available from: https://www.ncbi.nlm.nih.gov/books/NBK233371/

2. Blankenstein TN, Gibson LM, Claydon MA. Is intramuscular morphine satisfying frontline medical personnels’ requirement for battlefield analgesia in Helmand Province, Afghanistan? A questionnaire study. Br J Pain. 2015;9(2):115–121. DOI:10.1177/2049463714535563

3. Epstein TG, Liss GM, Berendts KM, Bernstein DI. AAAAI/ACAAI Subcutaneous Immunotherapy Surveillance Study (2013-2017): Fatalities, Infections, Delayed Reactions, and Use of Epinephrine Autoinjectors. J Allergy Clin Immunol Pract [Internet]. 2019 Jul 1 [cited 2020 Jul 27];7(6):1996–2003.e1. Available from: https://pubmed.ncbi.nlm.nih.gov/30776526/

4. Epstein T, Liss GM, Murphy-Berendts KJ, Bernstein DI. Recent Trends in Fatalities, Waiting Times, and Use of Epinephrine Autoinjectors for Subcutaneous Allergen Immunotherapy (SCIT): AAAAI/ACAAI National Surveillance Study 2008-2016. J Allergy Clin Immunol. 2018 Feb ;141(2):AB401.

5. Campbell RL, Bellolio MF, Motosue MS, Sunga KL, Lohse CM, Rudis MI. Autoinjectors preferred for intramuscular epinephrine in anaphylaxis and allergic reactions. West J Emerg Med [Internet]. 2016 [cited 2020 Jul 27];17(6):775–81. Available from: /pmc/articles/PMC5102607/?report=abstract

6. Nelson SC, Wedgwood JTAHow much do soldiers know about the morphine they carry on operations? A questionnaire study of knowledge and understanding of the morphine autoinjector on Op HERRICK 17BMJ Military Health 2015;161:27–31.

7. Tariq RA, Vashisht R, Sinha A, et al. Medication Dispensing Errors And Prevention. [Updated 2021 Feb 16]. In: StatPearls [Internet]. Treasure Island (FL): StatPearls Publishing; 2021 Jan–. Available from: https://www.ncbi.nlm.nih.gov/books/NBK519065/

8. Vijayaraghavan R. Autoinjector device for rapid administration of drugs and antidotes in emergency situations and in mass casualty management. J Int Med Res. 2020;48(5):300060520926019. doi:10.1177/0300060520926019

9. Potera, Carol Misuse of Autoinjectors and Inhalers, AJN, American Journal of Nursing: March 2015 - Volume 115 - Issue 3 - p 17 doi: 10.1097/01.NAJ.0000461799.44904.d3

10. Vasileiou K, Barnett J, Thorpe S, Young T. Characterising and justifying sample size sufficiency in interview-based studies: systematic analysis of qualitative health research over a 15-year period. BMC Med Res Methodol. 2018 Nov 21;18(1):148. doi: 10.1186/s12874-018-0594-7. PMID: 30463515; PMCID: PMC6249736.

11. Roy, A., Geetha, R. V., Magesh, A., Vijayaraghavan, R., & Ravichandran, V. (2021). Autoinjector - A smart device for emergency cum personal therapy. Saudi pharmaceutical journal : SPJ : the official publication of the Saudi Pharmaceutical Society, 29(10), 1205–1215. https://doi.org/10.1016/j.jsps.2021.09.004

12. Weinhold, T., Del Zotto, M., Rochat, J. et al. Improving the safety of disposable auto-injection devices: a systematic review of use errors. AAPS Open 4, 7 (2018). https://doi.org/10.1186/s41120-018-0027-z

13. Chaby, L. E., Sheriff, M. J., Hirrlinger, A. M., & Braithwaite, V. A. (2015). Can we understand how developmental stress enhances performance under future threat with the Yerkes-Dodson law?. Communicative & integrative biology, 8(3), e1029689. https://doi.org/10.1080/19420889.2015.1029689

14. Makwana, S., Basu, B., Makasana, Y., & Dharamsi, A. (2011). Prefilled syringes: An innovation in parenteral packaging. International journal of pharmaceutical investigation, 1(4), 200–206. https://doi.org/10.4103/2230-973X.93004

